# Muse 2 forehead photoplethysmography agrees with Polar H10 for heart rate but overestimates ultra-short RMSSD

**DOI:** 10.64898/2026.10.05.26364782

**Authors:** I Putu Adiartha Griadhi, Ida Ayu Astiti Suadnyana, Nila Wahyuni

**Author notes:** **Correspondence:** I Putu Adiartha Griadhi, Department of Physiology, Faculty of Medicine, Universitas Udayana, Denpasar, Bali, Indonesia.

## Abstract

The Muse 2 EEG headset also records forehead photoplethysmography (PPG). We tested whether its pulse intervals can replace Polar H10 RR intervals for ultra-short heart-rate variability in 28 seated recordings (60 s eyes open, then 60 s eyes closed), using fixed processing, no exclusions and eleven sensitivity analyses. Mean heart rate agreed almost perfectly (ICC ≥ 0.999; bias −0.22 and 0.00 beats min−1). SDNN agreement was good to excellent (ICC 0.86 and 0.91; bias +8.6 and +8.2 ms). RMSSD agreement was moderate (ICC 0.60): Muse exceeded Polar in at least 27 of 28 sessions (bias +19.2 and +18.5 ms; about 1.9-fold), and the ratio bias fell as RMSSD rose. Interval differences (SD 23 ms) implied about 16 ms of independent timing error per pulse, which reproduced the RMSSD bias; without ECG, sensor jitter could not be separated from variation in pulse transit time. Tracking of the eyes-closed change in ln RMSSD was not demonstrated (ρ = 0.35, 95% CI −0.07 to 0.67; detectable ρ ≥ 0.51). Muse 2 PPG can supply mean heart rate and, with a stated bias, SDNN; its ultra-short RMSSD is not interchangeable with Polar RMSSD and should be reported as pulse-rate variability.

## 1. Introduction

Heart-rate variability (HRV) derived from the electrocardiogram (ECG) is the reference approach for quantifying cardiac autonomic timing (Shaffer and Ginsberg, 2017; Task Force of the European Society of Cardiology and the North American Society of Pacing and Electrophysiology, 1996). Several time-domain indices, including the root mean square of successive differences (RMSSD) and the standard deviation of normal-to-normal intervals (SDNN), can be estimated from ultra-short recordings of one to a few minutes (Baek et al., 2015; Munoz et al., 2015). Chest-strap monitors such as the Polar H10 provide RR intervals that agree closely with laboratory ECG at rest (Schaffarczyk et al., 2022).

Photoplethysmography (PPG) senses the arterial pulse optically. The intervals it provides are pulse-to-pulse intervals, and their variability is pulse-rate variability (PRV) rather than HRV, because pulse arrival time differs from the ECG R wave and varies with vascular tone, respiration and blood pressure (Gil et al., 2010; Kantrowitz et al., 2025; Schäfer and Vagedes, 2013). Any error in locating the pulse feature is amplified in indices based on successive differences. If independent errors of standard deviation σ affect the timing of each pulse, RMSSD^2^ is inflated by approximately 6σ^2^, whereas mean HR is almost unaffected and SDNN is affected much less. This predicts near-perfect agreement with a reference RR series for mean HR, intermediate agreement for SDNN and the poorest agreement for RMSSD, with a positive RMSSD bias that is largest, in relative terms, when true variability is low.

The Muse 2 (InteraXon, Toronto, Canada) is a consumer four-channel EEG headband with a forehead PPG sensor sampled at 64 Hz. Simultaneous EEG and pulse signals make it attractive for studies of brain–heart interaction. The forehead is a demanding site: blinking and frontalis activity cause motion artifact, contact pressure is set by the headband, and 64 Hz sampling quantises pulse timing to 15.6 ms. Skin pigmentation, activity and device design are known sources of error in wearable optical sensors (Bent et al., 2020). Validation should use a criterion signal and agreement statistics rather than correlation alone (Bland and Altman, 1986, 1999; Kottner et al., 2011; Mühlen et al., 2021).

We searched PubMed and Google Scholar in September 2026 (terms: Muse 2 AND (PPG OR photoplethysmography) AND (HRV OR RMSSD OR Polar OR ECG OR validation)). The search did not identify a peer-reviewed concurrent-validity study of Muse 2 forehead pulse intervals against an ECG or chest-strap RR series. Muse 2 multimodal protocols have used the optical stream to derive heart rate without reporting beat-to-beat RMSSD agreement against ECG or a chest strap.

We tested the concurrent validity of Muse 2 pulse intervals against Polar H10 RR intervals for mean HR, RMSSD and SDNN over 60 s windows of seated eyes-open and eyes-closed rest. We asked whether any disagreement is consistent with pulse-timing error, which other explanations remain open, whether commonly proposed remedies remove it, and whether the change in RMSSD between the two conditions is tracked. The processing pipeline was fixed for all sessions, with no selection based on the reference signal.

## 2. Materials and Methods

### 2.1. Design, participants and ethics

This was a single-session concurrent-validity study with the Polar H10 as the reference. Twenty-eight hospitality employees (P01–P28; one recording per participant) were recorded on 7 and 8 September 2026. Participants were 23 women and 5 men, aged 30.9 ± 7.8 years (range 20–48). Height was 160.4 ± 8.9 cm, body mass 60.4 ± 12.8 kg, and body-mass index 23.3 ± 3.7 kg m−2 (range 17.5–31.2). Skin phototype was not recorded. Participants sat quietly and were able to wear both devices for about four minutes. The study was approved by the Research Ethics Committee of Universitas Dhyana Pura (protocol No. 006821/KEP Universitas Dhyana Pura/2026, 25 August 2026). All participants provided written informed consent. The study complied with the Declaration of Helsinki.

### 2.2. Devices and protocol

The Muse 2 was worn on the forehead according to the manufacturer’s instructions. Its PPG stream (three optical channels exported as PPG_1–PPG_3; PPG_2, the infrared channel, was used throughout) was sampled at 64 Hz. EEG and accelerometer streams were recorded simultaneously but are not analyzed here; the EEG is the subject of a companion report. Data were streamed with Lab Streaming Layer and stored as XDF files, with comma-separated exports for 26 sessions. For two sessions (P04 and P07) only the XDF file was available; the PPG stream was decoded with a custom reader because the sample-count field in these files is non-standard. The Polar H10 chest strap provided RR intervals with 1 ms resolution, saved as text files without per-beat timestamps. Raw ECG was not recorded.

After a countdown of three, the start buttons of both devices were pressed together. Participants sat with the eyes open for 60 s and then closed the eyes for about 180 s. The order was fixed, so condition and time on task are confounded. Muse recordings lasted 248–264 s (median 251 s) and Polar recordings 127–264 s (median 182 s). Only the first 120 s were compared, because the shortest Polar recording lasted 127 s: eyes open (EO, 0– 60 s), eyes closed (EC, 60–120 s) and a sensitivity window (EC-clean, 70–120 s) that discarded the first 10 s after eye closure. Windows were placed as recorded.

### 2.3. PPG processing

Zero-valued samples were replaced by linear interpolation and the signal was band-pass filtered (zero-phase Butterworth, third order, 0.6–3.5 Hz). Systolic peaks were detected with a minimum inter-peak distance corresponding to 160 beats min−1 and a prominence of 1.4826 times the median absolute deviation of the filtered signal. Pulse-to-pulse intervals were differences of successive peak times. The infrared channel (PPG_2) with positive polarity was used for all sessions. An earlier exploratory version chose channel and polarity per session by agreement with the reference; that procedure was abandoned and is reproduced only as sensitivity analysis S5.

### 2.4. Polar processing, artifact handling and indices

Polar beat times were reconstructed by cumulative summation of RR intervals from the start of the file. The same rule was applied to both devices: intervals outside 300–2000 ms, or differing by more than 20% from the immediately preceding interval, were rejected. Mean HR was 60 000 divided by the mean NN interval, SDNN was the sample standard deviation of NN intervals, and RMSSD was the root mean square of successive differences computed only between consecutive valid intervals. The artifact fraction was the proportion of intervals rejected in a window.

### 2.5. Clock alignment

Windows were placed on the time since the start of each recording. The Polar series was related to the Muse timeline by the difference of the recording-start timestamps (for P04 and P07, which lack metadata, the median difference). Pairing of individual heart beats showed that Muse time was earlier than Polar time by a median of 3.7 s. Of this, 0.9 s is the start-timestamp difference and a further 2.8 s (SD 0.8 s) is unresolved; it is not a pulse transit time (beat-to-pulse delay ≈ 0.2 s). Because 60 s indices are insensitive to a shift of this size, the nominal windows were kept for the primary analysis. Sensitivity analyses used per-session beat-level offsets (S10) and the start timestamps plus one constant lag (S11). The beat-level analysis used the offset that minimised the RMSE of matched consecutive pairs.

### 2.6. Quality control and statistics

The primary analysis included all 28 sessions. A reference-free quality-control subset comprised sessions with a Muse artifact fraction ≤ 0.10 over 0–120 s (n = 25). Agreement (Muse − Polar) was quantified with ICC(2,1) (Shrout and Fleiss, 1979) and a percentile bootstrap 95% CI (3000 resamples), Lin’s concordance correlation coefficient (CCC) (Lin, 1989), Pearson’s r, Spearman’s ρ, MAPE, and Bland–Altman bias and 95% limits of agreement (LoA). ICC values were described as poor (< 0.5), moderate (0.5–0.75), good (0.75–0.9) or excellent (> 0.9) (Koo and Li, 2016). Proportional bias was examined by regressing the difference on the mean. RMSSD was also analyzed after natural-log transformation, in which the bias is a ratio. For mean HR we considered agreement adequate if the absolute bias was below 1 beat min−1 and the LoA lay within ±5 beats min−1. Tracking of the change from eyes open to eyes closed used Spearman ρ of Δ ln RMSSD. No sample-size calculation was performed; precision is expressed by confidence intervals. The processing pipeline, windows and agreement criteria were fixed after the data had been inspected, before the final analysis: an earlier exploratory version selected the PPG channel and polarity per session by agreement with the reference and was abandoned (S5). The study was not preregistered. Reporting followed GRRAS and HRV recommendations (Kottner et al., 2011; Laborde et al., 2017).

### 2.7. Timing-noise model

Assume each observed pulse time equals the true pulse time plus an independent error e_i with variance σ^2^. The error in one pulse-to-pulse interval is e_{i+1} − e_i and therefore has variance 2σ^2^. RMSSD is the root-mean-square of successive interval differences. Consecutive intervals share one pulse time, so the error in a successive difference is (e_{i+2} − e_{i+1}) − (e_{i+1} − e_i) = e_{i+2} − 2e_{i+1} + e_i, which has variance 6σ^2^ if the e_i are independent and identically distributed. Plotting Muse RMSSD^2^ against Polar RMSSD^2^ should therefore have slope near 1 and intercept near 6σ^2^. σ was also estimated as the within-session SD of Muse − Polar interval differences divided by 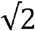. The model treats Polar intervals as the reference series for this comparison; it does not assume that the Polar H10 is free of all timing error. The model also cannot tell measurement error from genuine beat-to-beat variation in pulse arrival time: both enter e_i. Raw ECG was not recorded, so pulse transit time could not be measured, and σ is an effective value that combines the two.

### 2.8. Sensitivity analyses

Eleven variants were run:

- S1, exclusion of the two participants with Polar RMSSD > 80 ms;
- S2, the QC subset;
- S3, RMSSD computed across removed intervals;
- S4, a local-median artifact rule;
- S5, the alternate PPG channel for one session;
- S6, clock offset shifted by ±2 s;
- S7, peak prominence of 0.5 and 1.5 times the MAD;
- S8, parabolic peak interpolation;
- S9, a 0.5–8 Hz pass-band;
- S10, per-session beat-level offsets;
- S11, start timestamps plus one constant lag.

### 2.9. Use of generative AI

Generative AI (Claude, Anthropic) assisted with writing the analysis code: the original analysis script (`muse_polar_analysis.py`), including the eleven sensitivity analyses, and the scripts that summarize the beat-level and clock-alignment outputs, draw the figures and export the shared beat-level data. It was also used to re-run the analysis on the raw recordings to confirm that the script reproduces every table, to check references against Crossref and to cross-check every reported number against the analysis tables. The authors reviewed the code, its output and the text and take full responsibility for them.

## 3. Results

### 3.1. Data completeness

All 28 paired recordings entered the primary analysis. Muse artifact fraction over 0–120 s was ≤ 0.10 in 25 sessions. Polar covered EO and EC in every session.

### 3.2. Mean heart rate

Mean HR agreed almost perfectly (Table 1, Fig. 1). ICC(2,1) was 0.999 (0.997–1.000) with the eyes open and 1.000 (0.999–1.000) with the eyes closed. Bias was −0.22 beats min−1 (−0.38 to −0.05) and 0.00 (−0.12 to 0.11). Limits of agreement were −1.1 to 0.6 and −0.6 to 0.6 beats min−1. MAPE was 0.48% and 0.28%. Both windows met the HR criterion stated in Section 2.6.

**Table 1.** Primary agreement (n = 28). ICC is ICC(2,1) with bootstrap 95% CI. Bias is Muse − Polar.

| Metric / window | n | ICC [95% CI] | Bias [95% CI] | 95% LoA |
| --- | --- | --- | --- | --- |
| HR EO, beats min<sup>-1</sup> | 28 | 0.999 [0.997, 1.000] | -0.22 [-0.38, -0.05] | -1.1 to 0.6 |
| HR EC, beats min<sup>-1</sup> | 28 | 1.000 [0.999, 1.000] | 0.00 [-0.12, 0.11] | -0.6 to 0.6 |
| RMSSD EO, ms | 28 | 0.60 [0.34, 0.77] | +19.2 [14.9, 23.4] | -2.2 to 40.6 |
| RMSSD EC, ms | 28 | 0.60 [0.38, 0.75] | +18.5 [14.0, 23.0] | -4.2 to 41.2 |
| SDNN EO, ms | 28 | 0.86 [0.73, 0.95] | +8.6 [4.9, 12.4] | -10.4 to 27.7 |
| SDNN EC, ms | 28 | 0.91 [0.79, 0.95] | +8.2 [5.8, 10.5] | -3.6 to 19.9 |

**Fig. 1.**
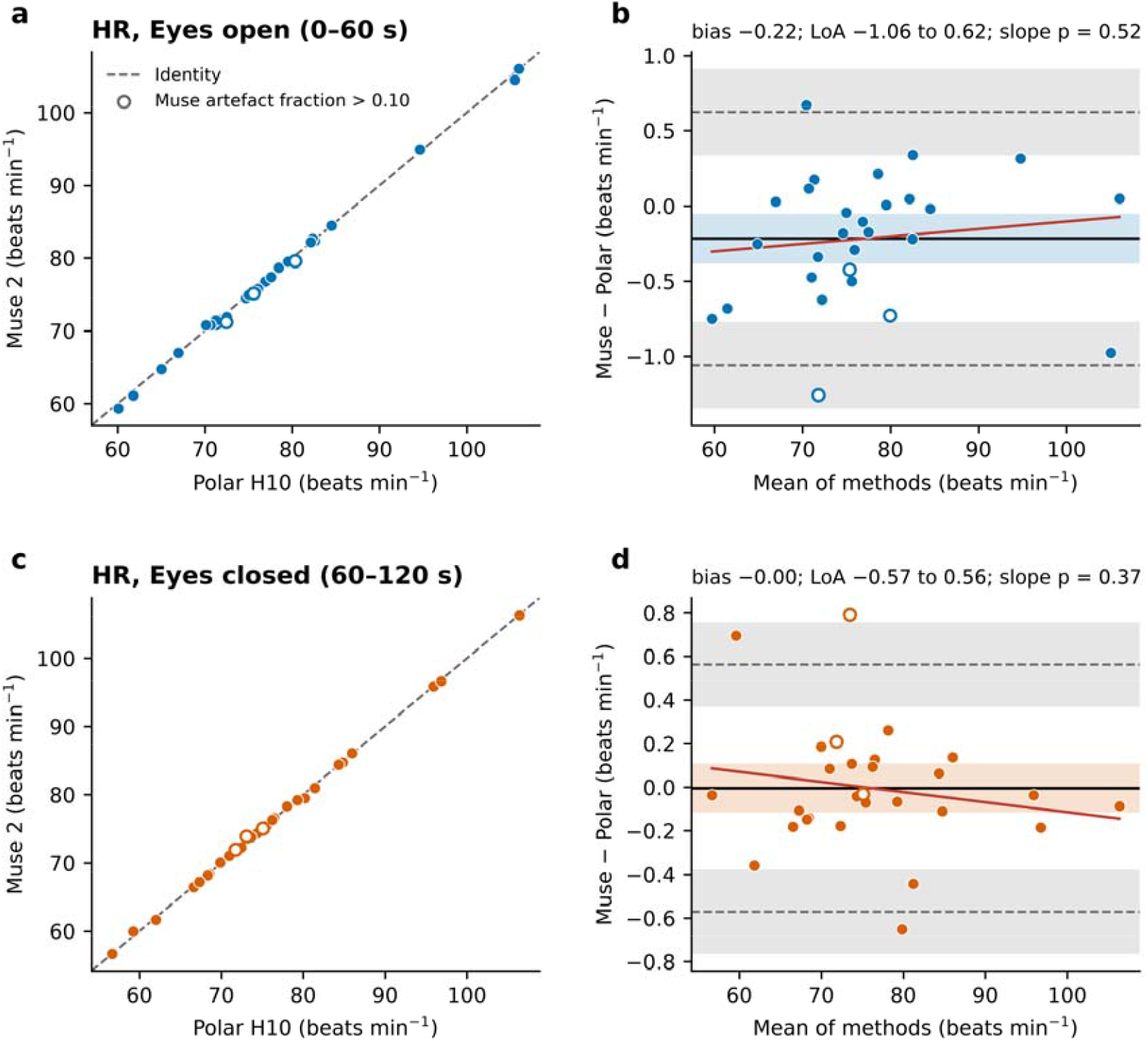
Agreement of session mean heart rate. Identity plots (a, c) and Bland–Altman plots (b, d) for eyes open and eyes closed. Open circles: Muse artifact fraction > 0.10.

### 3.3. RMSSD

RMSSD agreement was moderate (Table 1, Fig. 2). ICC(2,1) was 0.60 (0.34–0.77) eyes open and 0.60 (0.38–0.75) eyes closed. Muse exceeded Polar in 27 of 28 sessions eyes open and in all 28 eyes closed. Bias was +19.2 ms (14.9 to 23.4) and +18.5 ms (14.0 to 23.0), about 1.9-fold on the log scale (ratio 1.88 and 1.89). Limits of agreement were −2.2 to 40.6 ms and −4.2 to 41.2 ms. The log-scale slope of the difference on the mean was −0.75 and −0.76 (p < 0.001): the relative overestimate was largest when Polar RMSSD was low. Excluding the two participants with Polar RMSSD > 80 ms lowered ICC to 0.43 and 0.51.

**Fig. 2.**
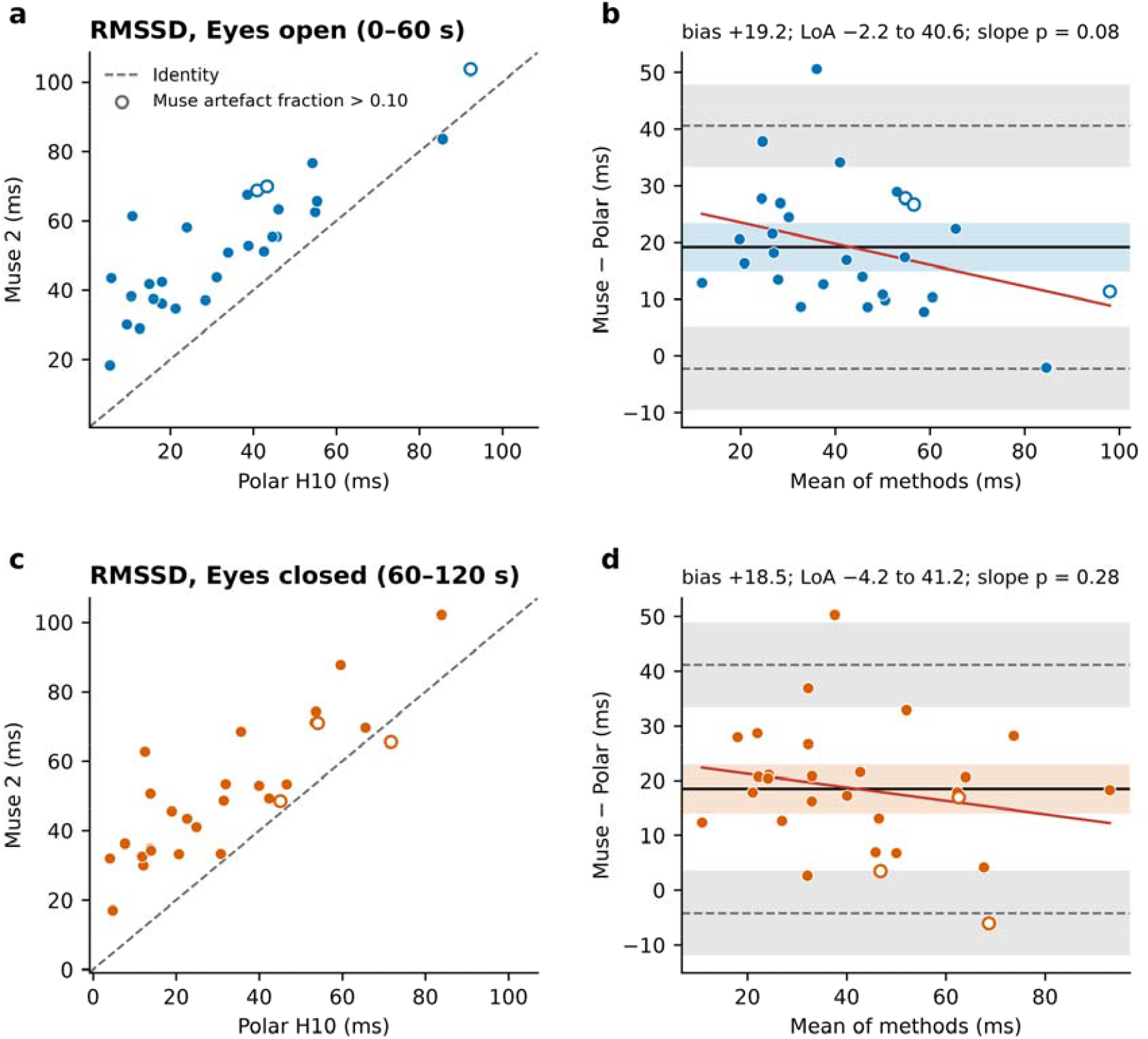
Agreement of 60 s RMSSD. Identity plots (a, c) and Bland–Altman plots (b, d). Muse values lie above the identity line in almost every session.

### 3.4. SDNN

SDNN agreement with the Polar H10 was good with the eyes open and excellent with the eyes closed (Table 1, Fig. 3). ICC(2,1) was 0.86 (0.73–0.95) and 0.91 (0.79–0.95). Bias was +8.6 ms (4.9 to 12.4) and +8.2 ms (5.8 to 10.5). Limits of agreement were −10.4 to 27.7 ms and −3.6 to 19.9 ms. There was no consistent proportional bias.

**Fig. 3.**
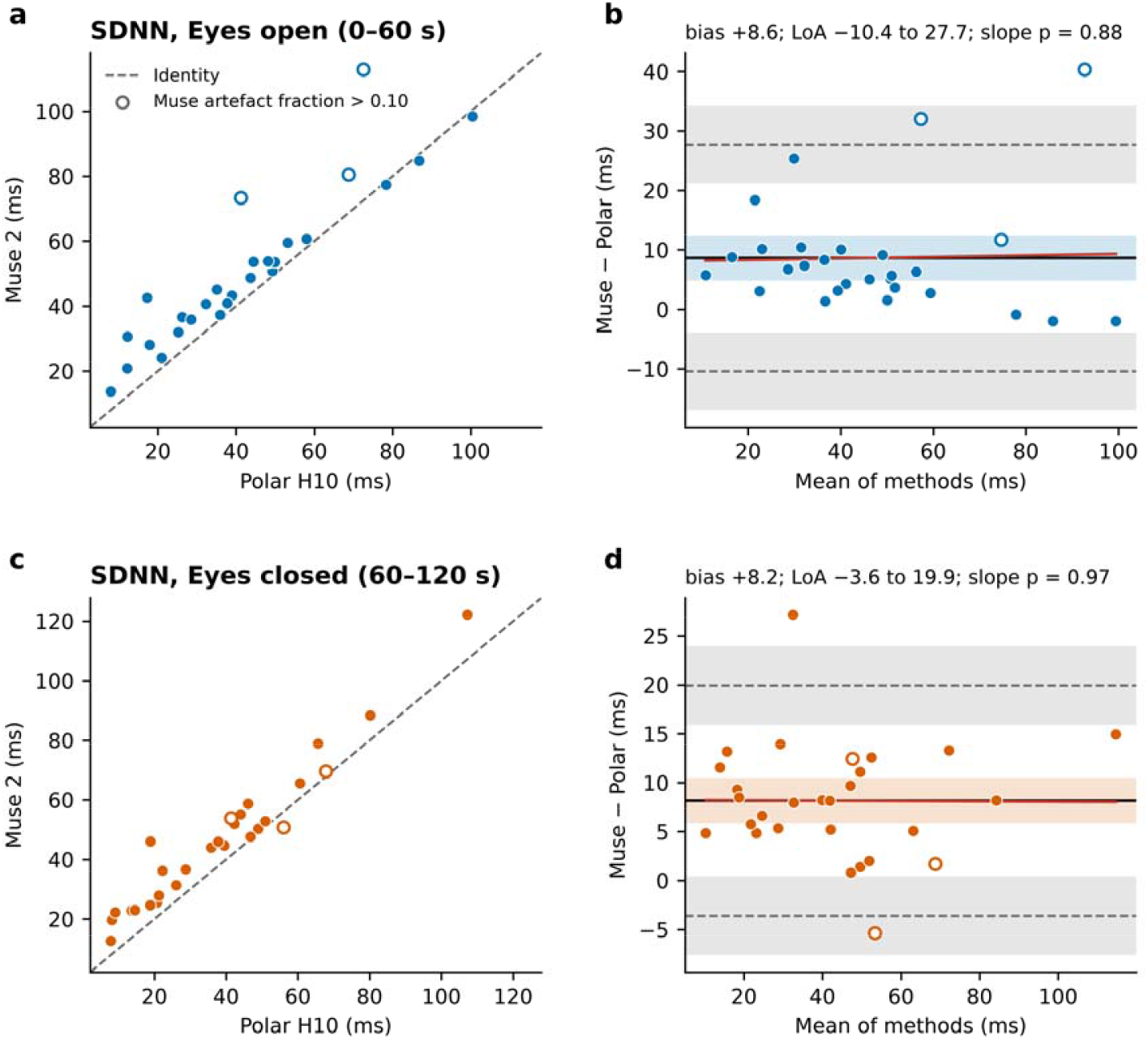
Agreement of 60 s SDNN. Identity plots (a, c) and Bland–Altman plots (b, d).

### 3.5. Timing-error model

The beat-level comparison paired each Muse pulse with a Polar beat after choosing the clock offset that minimised the interval error, so it describes agreement under the most favourable alignment; σ is a lower bound for the effective timing error. Median within-session SD of Muse − Polar interval differences was 23.4 ms (IQR 19.3–29.5), corresponding to σ ≈ 16.5 ms per pulse. Muse RMSSD^2^ versus Polar RMSSD^2^ had slope 0.98 (eyes open) and 1.12 (eyes closed) and intercepts that implied σ of 16.1 and 14.7 ms, matching the beat-level estimate. Adding 6σ^2^ with σ = 16 ms to the identity line described the cloud (Fig. 4b). Parabolic interpolation reduced interval-error SD only from 23.4 to 22.5 ms and changed RMSSD bias by less than 2 ms. Widening the pass-band to 0.5–8 Hz increased interval-error SD to 36.6 ms (Supplementary Table S2) and lowered RMSSD ICC to 0.39 and 0.36.

**Fig. 4.**
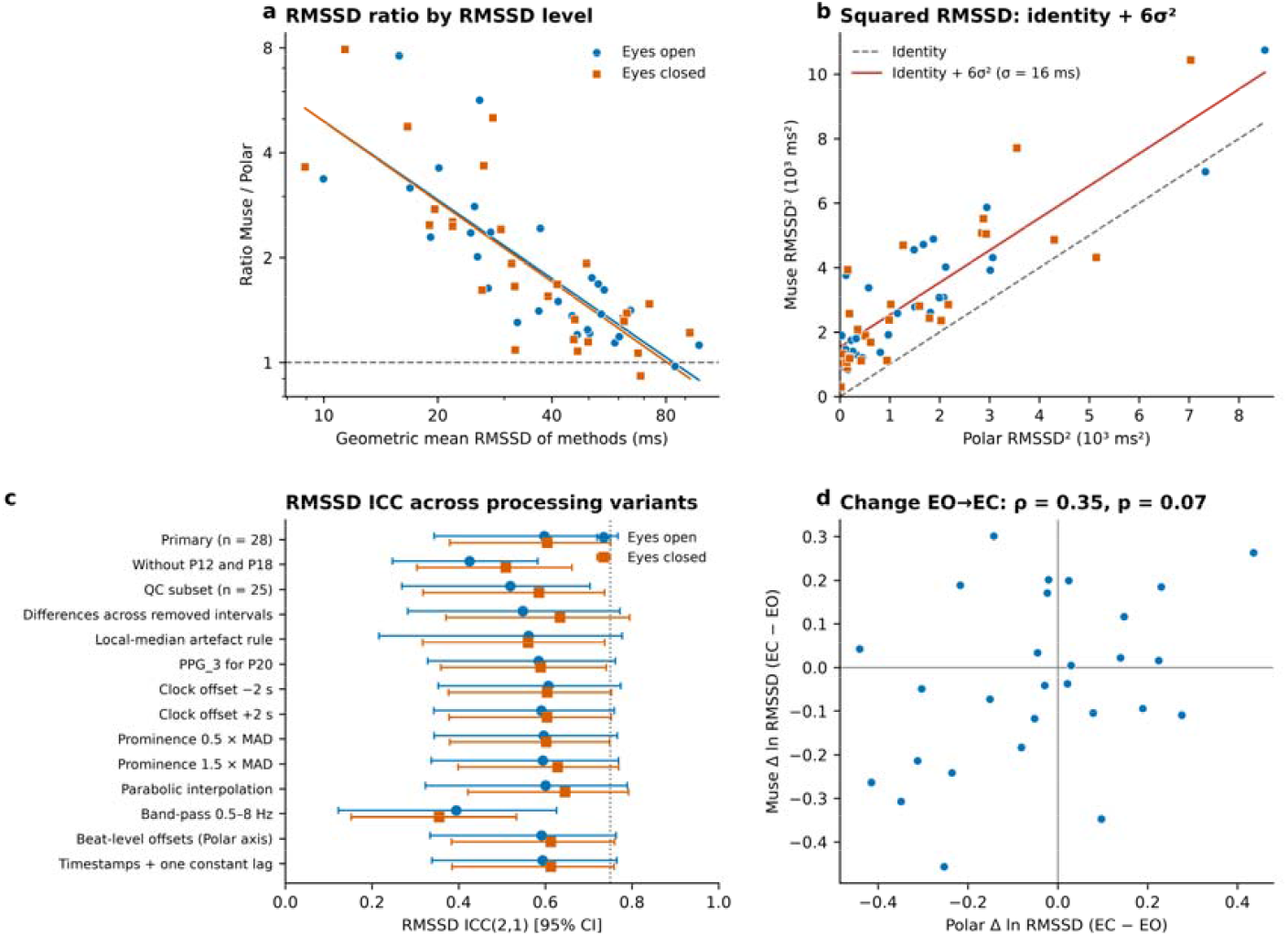
Source of the RMSSD bias. (a) Muse/Polar RMSSD ratio falls as the geometric mean rises. (b) Muse RMSSD^2^ versus Polar RMSSD^2^ with the identity + 6σ^2^ line (σ = 16 ms). (c) RMSSD ICC across processing variants. (d) Change in ln RMSSD: tracking was not demonstrated (ρ = 0.35).

### 3.6. Change tracking and sensitivity

Spearman ρ between Muse and Polar Δ ln RMSSD (EC − EO) was 0.35 (p = 0.07). Directional agreement was 61% (17 of 28). The bootstrap 95% CI for ρ was −0.07 to 0.67, and with 28 participants only correlations of about 0.51 or larger would have been detected with 80% power. The data are therefore compatible with anything from no tracking to moderately good tracking: this is insufficient evidence of reliable change tracking, not a demonstration that tracking is impossible. In contrast, the change in mean HR was tracked closely (r = 0.98). Across the eleven sensitivity analyses, RMSSD ICC stayed between 0.52 and 0.65 (Fig. 4c; Supplementary Table S1), with two exceptions. It fell to 0.43 and 0.51 when the two participants with Polar RMSSD > 80 ms were excluded, and to 0.39 and 0.36 when high-frequency noise was admitted. The bias stayed between +17 and +23 ms except with the wider pass-band. QC restriction (n = 25) and beat-level clock realignment did not remove the +19 ms bias.

## 4. Discussion

### 4.1. Principal findings

Forehead PPG from the Muse 2 showed near-equivalent session mean heart rate to the Polar H10 during seated rest. SDNN agreement was good to excellent, with a positive bias of about 8–9 ms. Ultra-short RMSSD is not interchangeable with the Polar H10: the headset overestimates by about 19 ms, or 1.9-fold, and the relative error is worst where vagal markers are most often interpreted, in people with low variability. The pattern is consistent with about 16 ms of effective timing error per pulse. This error may come from the sensor, from physiological variation in pulse transit time, or from both (Section 4.2).

### 4.2. Possible sources of the RMSSD bias

Mean rate averages away independent timing error. Successive differences amplify it. The data fit a model with about 16 ms of independent timing error per pulse, but two sources can produce such an error and this design cannot separate them. The first is measurement jitter: a 64 Hz sensor quantises peaks to 15.6 ms (uniform quantisation alone corresponds to an SD of about 4.5 ms), and forehead motion, frontalis activity and changes in pulse shape can shift the detected peak. We did not isolate these as separate experimental factors. The second is physiological. Pulse arrival time varies from beat to beat with blood pressure, vascular tone and respiration, so PRV can differ from HRV even with a perfect sensor (Gil et al., 2010; Schäfer and Vagedes, 2013). Raw ECG and a second pulse site were not recorded, so pulse transit time could not be measured, and the 16 ms value should be read as the combined effect of both sources. The model is therefore a working hypothesis, not a demonstrated noise floor of the device. Interpolation of the peak position changed the bias by less than 2 ms, so simple sampling granularity is not a sufficient explanation. A wider pass-band made agreement worse, which points to at least some high-frequency measurement noise, but it does not rule out a physiological contribution. A study with simultaneous ECG, or with PPG at two sites, could separate the two. The fixed eyes-open-first order prevents separation of eye opening from time on task; that limitation applies to the change analysis, not to the within-window Muse–Polar comparison.

### 4.3 Practical implications

Session mean HR from the Muse 2 agreed with the Polar H10 closely enough for seated-rest monitoring in this sample. SDNN can be used with a stated positive bias of about 8–9 ms. Absolute RMSSD should not be treated as interchangeable with Polar RMSSD and is least trustworthy when Polar RMSSD is low. Muse-derived indices should be reported as PRV, not HRV (Kantrowitz et al., 2025). The present data give insufficient evidence that Muse tracks within-person change in RMSSD (ρ = 0.35; 61% directional agreement); they do not prove that tracking is impossible. Mean HR change was tracked. Studies that combine Muse EEG with autonomic variables should treat a null RMSSD finding as measurement-limited relative to this reference device.

### 4.4. Methodological lessons

Selecting channel, polarity or sessions with the help of the reference inflates agreement and was avoided. RMSSD must be computed from truly successive intervals. A fixed 20% rejection rule can remove legitimate respiratory sinus arrhythmia. Adaptive rules based on the local distribution of successive differences have been proposed (Lipponen and Tarvainen, 2019); a local-median rule gave similar conclusions here (S4).

### 4.5. Limitations

The sample was modest and single-session, without a sample-size calculation, and the analysis choices were fixed after the data had been inspected. Skin phototype was not recorded. The protocol was 2 min of seated rest with a fixed order and does not apply to movement, exercise or the standard 5 min recording. The reference device was a consumer chest strap, not a 12-lead ECG; raw ECG was not recorded, which also prevented measurement of pulse transit time. Conclusions concern Muse 2 versus Polar H10, not Muse 2 versus clinical ECG. The two clocks differed by 3.7 s of partly unresolved origin; 60 s indices and S10–S11 were insensitive to that shift. One device model, one 64 Hz pipeline and one artifact rule were tested. One recording per participant precludes test–retest reliability.

### 4.6. Future work

Higher-rate or multi-wavelength timing, template-based pulse features, 5 min recordings, skin phototype, posture, and repeated-measures designs with a sufficient RMSSD signal-to-noise ratio remain to be tested. Agreement during movement is also untested. Whole-body exercise such as moderate-intensity Legong dance training, which improves physical fitness (Griadhi et al., 2021), would add motion artifacts at the forehead and is a relevant setting for such a test.

## 5. Conclusion

During 2 min of seated rest, Muse 2 forehead PPG showed near-equivalent session mean heart rate to the Polar H10 (bias below 0.3 beats min−1) and high SDNN agreement with a positive bias of about +8 to +9 ms. Its RMSSD was higher than the Polar value in at least 27 of 28 participants (bias about +19 ms, about 1.9-fold), with moderate agreement. The bias is consistent with about 16 ms of effective timing error per pulse, from sensor jitter, physiological variation in pulse transit time, or both. Interpolation does not remove this error. The data give insufficient evidence that the change in RMSSD between conditions was tracked reliably. Muse RMSSD should be reported as pulse-rate variability, not as interchangeable with Polar RMSSD.

## Data availability statement

De-identified per-session indices, beat-level interval series (Muse pulse intervals and Polar RR intervals) and the aggregate tables that reproduce all reported statistics are openly available on Zenodo under a CC BY 4.0 license (Griadhi et al., 2026). Sessions are coded P01–P28 and contain no identifying information. The complete analysis code, including the eleven sensitivity analyses, is available in the same record (Griadhi et al., 2026) under the MIT license, with package versions fixed in `requirements.txt`.

## Ethics statement

The study was approved by the Research Ethics Committee of Universitas Dhyana Pura (protocol No. 006821/KEP Universitas Dhyana Pura/2026, 25 August 2026) and complied with the Declaration of Helsinki. All participants provided written informed consent.

## Conflict of interest

The authors have no competing interests to declare that are relevant to the content of this article.

## Funding information

This research did not receive any specific grant from funding agencies in the public, commercial, or not-for-profit sectors.

## Author contributions

I Putu Adiartha Griadhi: Conceptualization, Methodology, Software, Formal analysis, Investigation, Data curation, Writing – original draft, Visualization. Ida Ayu Astiti Suadnyana: Investigation, Writing – review and editing. Nila Wahyuni: Investigation, Writing – review and editing. All authors approved the submitted version.

## Acknowledgments

The authors thank the participants who sat for the dual-device recordings. Generative AI (Claude, Anthropic) assisted with drafting and editing the manuscript text; its use in the analysis is described in Section 2.9.

## Notes

### Competing Interest Statement

The authors have declared no competing interest.

### Author Declarations

Research Ethics Committee of Universitas Dhyana Pura gave ethical approval for this work

