## Supplemental Data 1 for "Muse 2 forehead photoplethysmography agrees with Polar H10 for heart rate but overestimates ultra-short RMSSD"

### Supplementary Table S1. RMSSD agreement across sensitivity analyses

ICC(2,1) with bootstrap 95% CI and bias (Muse − Polar, ms) for 60-s RMSSD; S0 is the primary analysis (Methods, Sensitivity analyses).

| Analysis | Window | n | ICC [95% CI] | Bias, ms [95% CI] |
| --- | --- | --- | --- | --- |
| S0 primary (n=28) | EO | 28 | 0.60 [0.34, 0.77] | +19.2 [14.9, 23.4] |
| S0 primary (n=28) | EC | 28 | 0.60 [0.38, 0.75] | +18.5 [14.0, 23.0] |
| S1 without P12 and P18 (Polar RMSSD > 80 ms) | EO | 26 | 0.43 [0.25, 0.58] | +20.3 [16.1, 24.5] |
| S1 without P12 and P18 (Polar RMSSD > 80 ms) | EC | 26 | 0.51 [0.30, 0.66] | +19.4 [15.0, 23.9] |
| S2 QC subset | EO | 25 | 0.52 [0.27, 0.70] | +18.8 [14.2, 23.5] |
| S2 QC subset | EC | 25 | 0.59 [0.32, 0.74] | +20.1 [15.7, 24.5] |
| S3 old RMSSD (differences across removed intervals) | EO | 28 | 0.55 [0.28, 0.77] | +22.0 [15.3, 28.8] |
| S3 old RMSSD (differences across removed intervals) | EC | 28 | 0.63 [0.37, 0.79] | +18.8 [14.2, 23.4] |
| S4 local-median artefact rule (25%) | EO | 28 | 0.56 [0.22, 0.78] | +23.2 [17.0, 29.4] |
| S4 local-median artefact rule (25%) | EC | 28 | 0.56 [0.32, 0.74] | +19.4 [12.2, 26.7] |
| S5 channel as in v1 (P20 = PPG_3) | EO | 28 | 0.58 [0.33, 0.76] | +19.5 [15.1, 23.8] |
| S5 channel as in v1 (P20 = PPG_3) | EC | 28 | 0.59 [0.36, 0.74] | +18.9 [14.3, 23.5] |
| S6 clock offset shifted -2 s | EO | 28 | 0.61 [0.35, 0.77] | +18.9 [14.7, 23.1] |
| S6 clock offset shifted -2 s | EC | 28 | 0.60 [0.38, 0.75] | +18.5 [14.0, 22.9] |
| S6 clock offset shifted +2 s | EO | 28 | 0.59 [0.34, 0.76] | +19.3 [15.2, 23.4] |
| S6 clock offset shifted +2 s | EC | 28 | 0.60 [0.38, 0.75] | +18.5 [14.1, 23.0] |
| S7 prominence 0.5xMAD | EO | 28 | 0.60 [0.34, 0.77] | +19.2 [15.0, 23.5] |
| S7 prominence 0.5xMAD | EC | 28 | 0.60 [0.38, 0.75] | +18.6 [14.1, 23.2] |
| S7 prominence 1.5xMAD | EO | 28 | 0.59 [0.34, 0.77] | +19.1 [14.7, 23.5] |
| S7 prominence 1.5xMAD | EC | 28 | 0.63 [0.40, 0.77] | +17.6 [13.3, 21.8] |
| S8 parabolic peak interpolation | EO | 28 | 0.60 [0.32, 0.79] | +18.5 [14.0, 23.1] |
| S8 parabolic peak interpolation | EC | 28 | 0.65 [0.42, 0.79] | +17.2 [13.0, 21.4] |
| S9 band-pass 0.5-8 Hz | EO | 28 | 0.39 [0.12, 0.63] | +25.8 [19.7, 31.9] |
| S9 band-pass 0.5-8 Hz | EC | 28 | 0.36 [0.15, 0.53] | +29.9 [24.0, 35.9] |
| S10 per-session beat-level offsets (windows on the Polar axis) | EO | 28 | 0.59 [0.33, 0.76] | +19.4 [15.0, 23.8] |
| S10 per-session beat-level offsets (windows on the Polar axis) | EC | 28 | 0.61 [0.38, 0.76] | +18.4 [14.1, 22.7] |
| S11 start timestamps plus one constant lag (windows on the Polar axis) | EO | 28 | 0.59 [0.34, 0.77] | +19.3 [14.9, 23.7] |
| S11 start timestamps plus one constant lag (windows on the Polar axis) | EC | 28 | 0.61 [0.39, 0.76] | +18.5 [14.2, 22.8] |

### Supplementary Table S2. Beat-level interval error

Within-session SD of Muse − Polar interval differences after beat-level alignment (median and IQR across 28 sessions) and the implied timing error per pulse (σ = SD/√2).

| Processing | Sessions | Interval-error SD, ms, median [IQR] | σ per pulse, ms |
| --- | --- | --- | --- |
| 0.6-3.5 Hz | 28 | 23.4 [19.3, 29.5] | 16.5 |
| 0.6-3.5 Hz + parabolic interpolation | 28 | 22.5 [18.4, 28.9] | 15.9 |
| 0.5-8 Hz | 28 | 36.6 [29.1, 39.1] | 25.9 |
